# Education and cardiovascular disease: a within-family Mendelian randomisation analysis

**DOI:** 10.1101/2025.07.02.25330770

**Authors:** Paul R Jones, Laxmi Bhatta, Laurence J Howe, María Fernanda Vinueza-Veloz, Neil M Davies, George Davey Smith, Øyvind E Næss, Ben M Brumpton

## Abstract

**Background:** Observational studies have consistently found educational inequalities in cardiovascular disease risk. Mendelian randomisation analyses have suggested a direct causal effect of education, however estimates may be biased by demography or dynastic effects. This study aimed to estimate the effects of educational attainment on cardiovascular disease risk and serum lipid concentrations before and after accounting for family structure.

**Methods:** This study included 28 448 siblings from the Trøndelag Health Study (HUNT), 27 040 siblings from UK Biobank, and >120 000 individuals from an international within-sibship genome-wide association study, predominantly of European ancestry. The exposure was educational attainment. The outcomes were cardiovascular disease risk and serum concentrations of low-density lipoprotein cholesterol, high-density lipoprotein cholesterol, and triglycerides. Standard and within-sibship Mendelian randomisation were used.

**Results:** In the summary data analysis, there was a 6% lower risk of cardiovascular disease (odds ratio 0.94, 95% confidence interval 0.92 to 0.96) for each additional standard deviation of educational attainment. This was consistent having accounted for family structure (odds ratio 0.96, 95% confidence interval 0.91 to 1.01). Educational attainment was also beneficially associated with each serum lipid concentration both before and after accounting for family structure. Results were broadly similar in the individual-level analysis.

**Conclusions:** There is a protective effect of educational attainment on cardiovascular disease risk and a beneficial effect on serum lipid concentrations not due to familial factors shared by siblings, suggesting that increasing education may be beneficial for cardiovascular health.

**KEY MESSAGES:** *Question:* Is the direct causal effect of education on cardiovascular disease (CVD) risk and CVD risk factors indicated by conventional Mendelian randomisation (MR) biased by demography or dynastic effects?

*Findings:* Consistent with conventional MR analyses, within-sibship MR indicated that higher educational attainment is protective CVD risk and beneficial for serum concentrations of low-density lipoprotein cholesterol, high-density lipoprotein cholesterol, and triglycerides.

*Importance:* Previously reported beneficial effects of educational attainment on CVD risk and serum lipid concentrations are likely causal and not due to bias from demographic or dynastic effects.

## BACKGROUND

The importance of educational attainment to socioeconomic inequalities in cardiovascular disease (CVD) is well recognised [1]. Observational studies have demonstrated consistent protective effects of higher educational attainment on CVD risk, which are already evident in middle age and contribute to substantial differences in lifetime risk [2,3]. Similarly, consistent favourable associations have been reported with CVD risk factors in high income countries [4–7]. Determining whether these beneficial associations of educational attainment are causal is challenging given the potential for confounding by early-life factors that potentially influence both educational attainment and CVD risk, such as higher childhood body mass index (BMI) [8,9].

Mendelian randomisation (MR) is an approach in which genetic variants associated with modifiable exposures of interest are used to strengthen evidence of causality on health outcomes (Explanatory box 1) [10,11]. Studies using MR methods have found evidence that educational attainment may be causal for risk of coronary heart disease (CHD) and ischaemic stroke [12,13], and a number of CVD risk factors [12,14,15]. Most MR studies to date have used samples of unrelated individuals and implicitly assume that the genetic variants used as proxies for educational attainment are inherited independent of confounding. However, in unrelated populations, random segregation of alleles is only an approximation, with evidence accumulating that GWASs for certain phenotypes also capture demographic or indirect genetic effects and can bias estimates of causal effects (Explanatory box 2) [16–19].

Family-based MR analysis can account for these possible alternative mechanisms to explain the relationship between educational attainment and CVD. Within a family, the offspring’s alleles are randomly inherited from their parents’ genotypes during meiosis. Any genetic differences between siblings are therefore random and independent of demographic (population stratification and assortative mating) and indirect genetic effects (many of which will be the same for siblings). Consequently, within-sibship analyses are robust to these mechanisms which can otherwise lead to biased estimates of the causal effects of education in MR analyses in unrelated samples. Until now, opportunities to conduct family-based MR were limited due to a paucity of genetic data for families. With the advent of national biobanks, sample sizes with adequate power are increasingly available [20].

In the MR context, binary and ordinal categorical exposures such as educational attainment are typically considered to have a latent continuous underlying liability that is normally distributed within the population [21,22]. This liability captures genetic, environmental and stochastic sources of variation but is observable in the expressed phenotype only once a threshold(s) has been crossed, such as stages of schooling. Here we estimate causal effects of liability to educational attainment on serum lipid concentrations and CVD risk with and without accounting for family structure to determine whether previously reported effects are robust to demographic or dynastic effects.

## Explanatory box 1

Figure adapted from Davey Smith and Hemani (2014)

### Mendelian randomisation

Mendelian randomisation is an approach to causal inference which leverages the random, independent allocation of parental alleles to offspring [10]. Genetic variants are used as an instrumental variable to determine the causal relationship between a modifiable disease risk factor and health outcomes [11,23]. Given that genetic germline variants are inherited randomly from parents at conception, their use as instrumental variables avoids issues of unmeasured confounding and reverse causation common with directly measured exposures in observational epidemiology [24]. In MR studies, instrumental variables are typically single nucleotide polymorphisms (SNPs) or polygenic scores (PGSs) identified as associated with an exposure through genome-wide association studies. To be a valid instrumental variable the genetic variant must satisfy three conditions: 1) it must be associated with the exposure of interest, 2) there should be no confounding of the instrumental variable and outcome, 3) any association with the outcome must be mediated via the exposure (Explanatory box 1 figure 1) [23].

**Explanatory box 1 figure 1.**
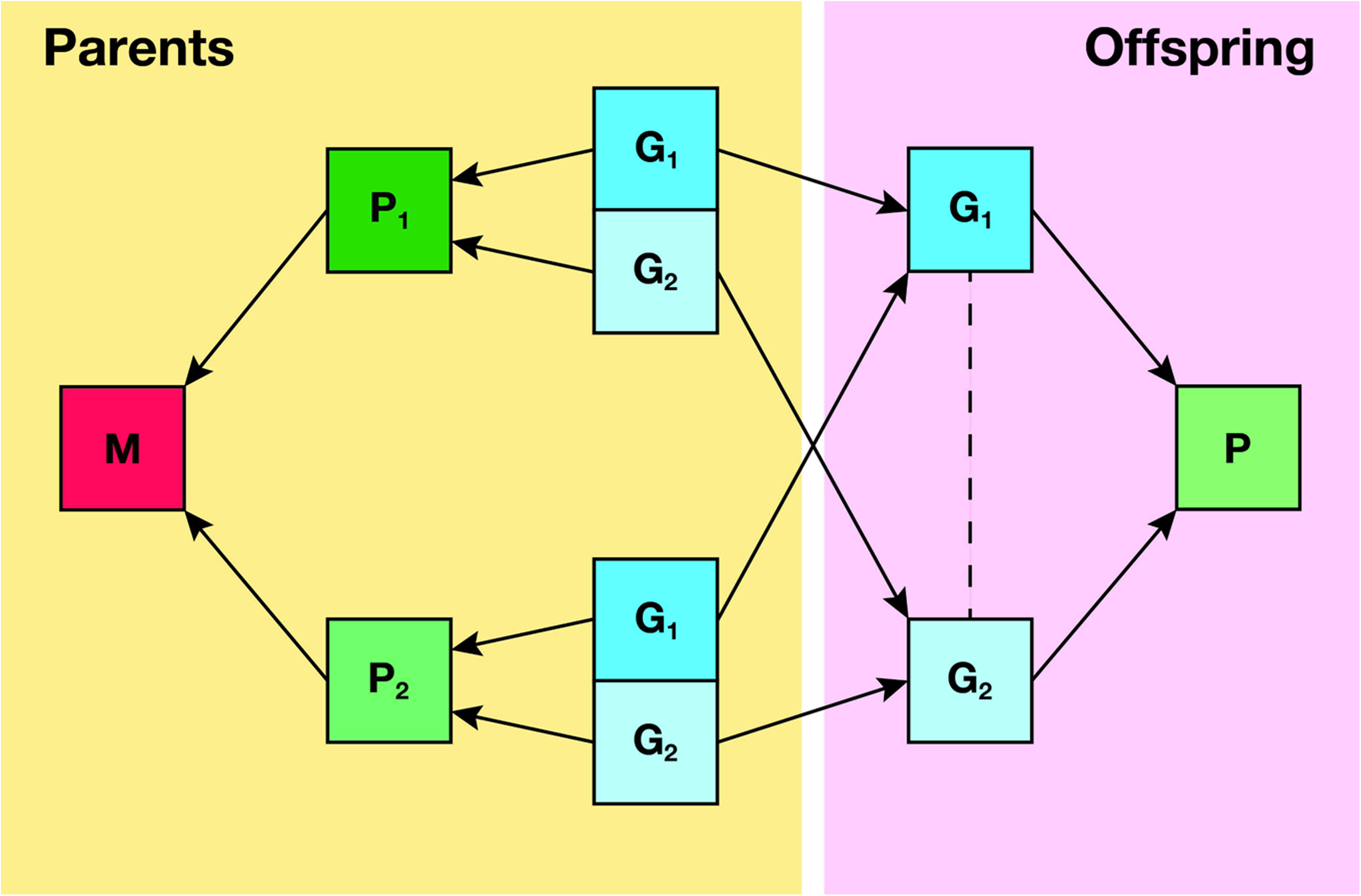
**Principals of Mendelian randomisation**

## Explanatory box 2

Figures adapted from Howe *et al.* (2022)

### Population stratification

Population stratification occurs when the distribution of a given allele(s) differs between groups within the same study population (Explanatory box 2 figure 1) [25]. This could produce a spurious association between genetic variants of an exposure and unrelated phenotypes which differ across populations. For example, in the UK Biobank there is evidence of geographical stratification of genetic variants associated with educational attainment that were not completely controlled by adjusting for genetic principal components [26]. If there are coincident geographic structures in certain disease outcomes, a MR analysis may incorrectly attribute a causal effect to education attainment.

**Explanatory box 2 figure 1.**
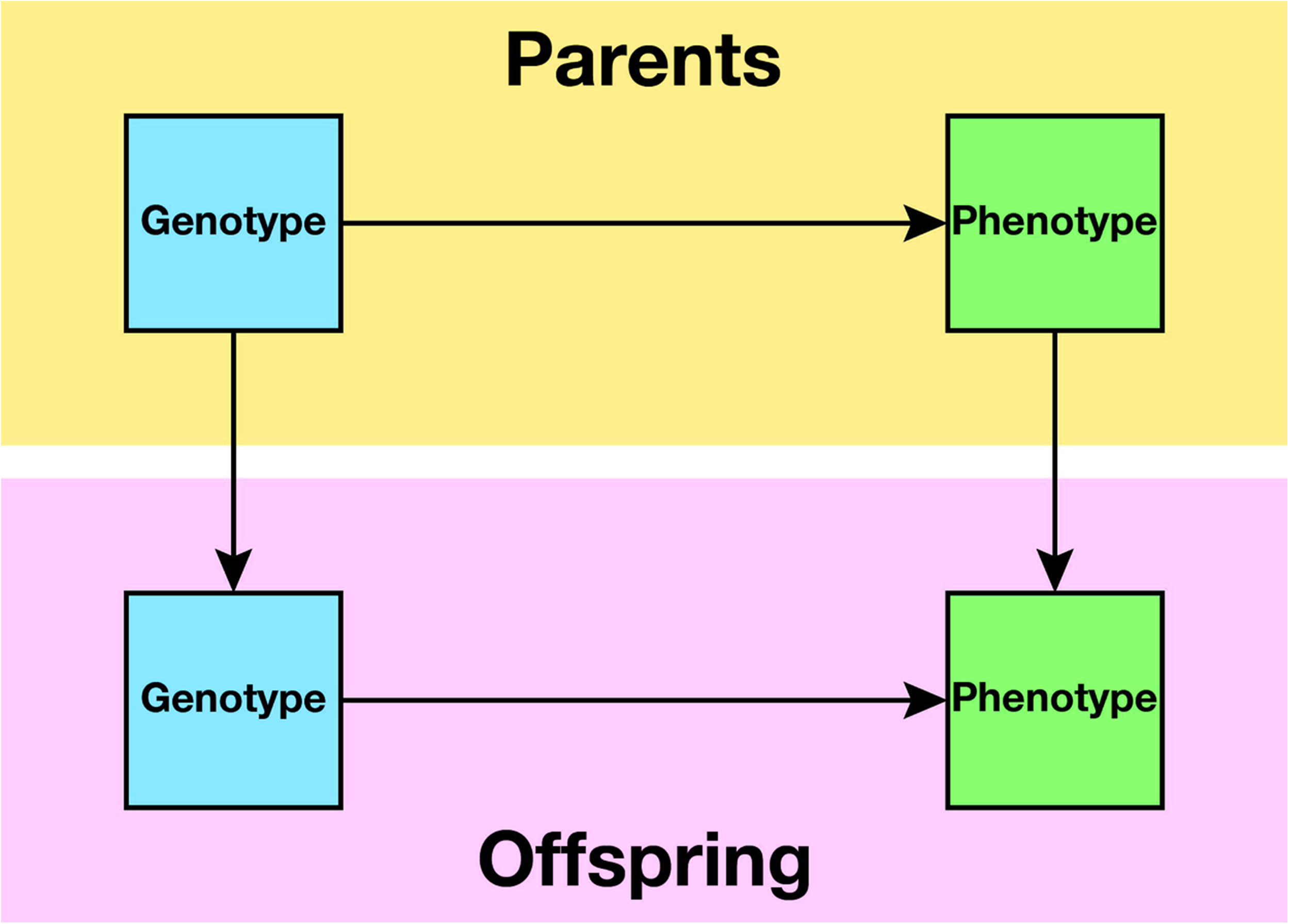
**Population stratification**

### Assortative mating

Assortative mating occurs when individuals with similar phenotypes, hence genotypes, are more likely to have offspring than would be expected under random mating (Explanatory box 2 figure 2) [27]. This can occur for a single trait, such as taller individuals selecting taller partners, or across traits, such as taller individuals selecting partners with higher earned income. Single trait assortative mating will produce bias in MR studies if the single phenotype is genetically correlated with both the exposure and outcome phenotypes, producing a correlation between the genetic variants for the exposure in one parent and variants for the outcome in the other parent [27]. This will likely result in overestimation of the causal effect of genetic traits on the exposure in offspring. Cross-trait assortative mating can induce associations between the genetic variants for exposure and outcome, biasing the association between exposure variants and outcome in offspring [28]. For cross-trait assortative mating, this bias can accumulate across subsequent generations [27].

**Explanatory box 2 figure 2.**
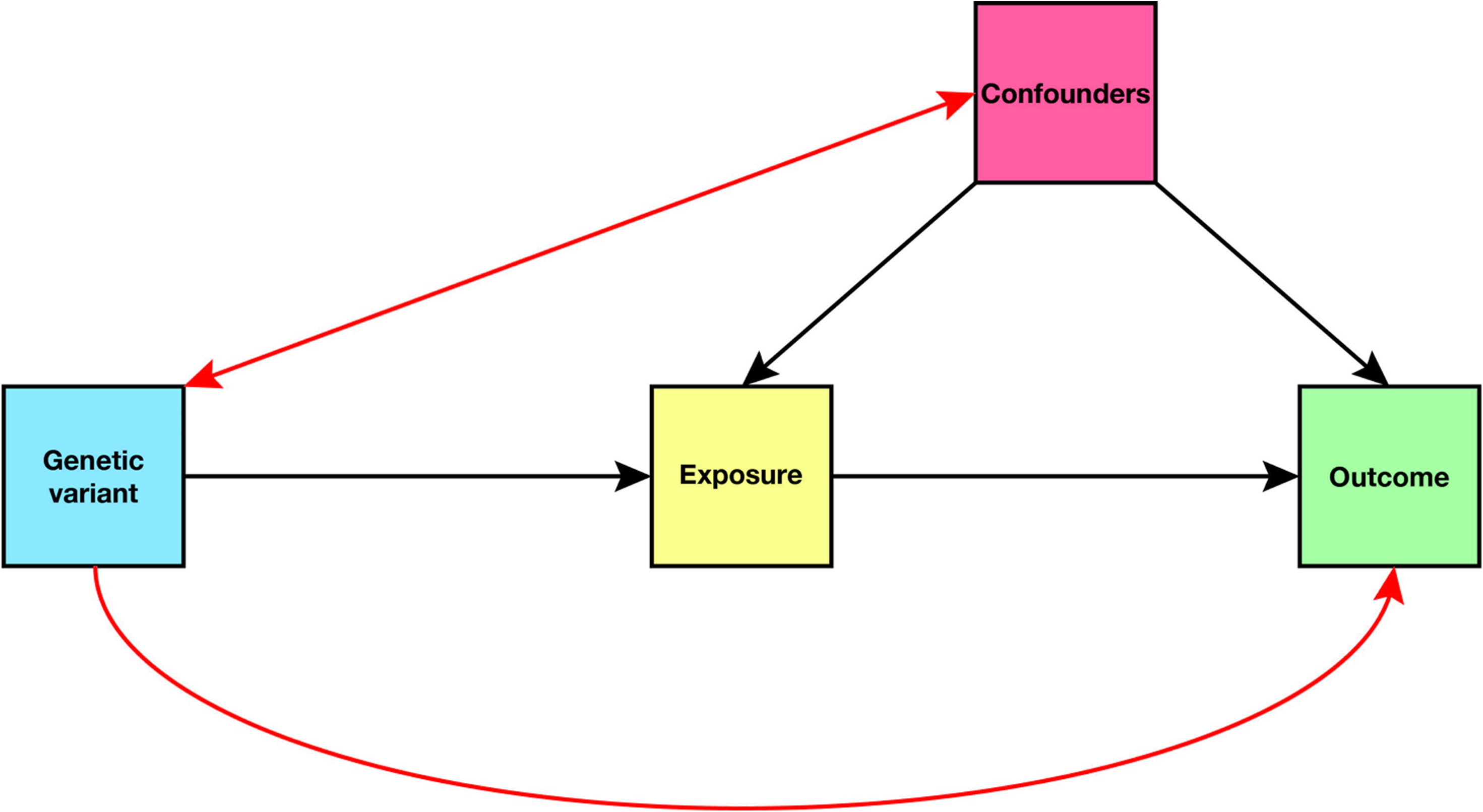
Assortative mating.

### Dynastic effects

Parental genotype directly affects offspring phenotype through genetic inheritance. By contrast, a dynastic effect (which is one example of an indirect genetic effect) occurs when the parental genotype affects the offspring phenotype indirectly through expression in the parental phenotype (Explanatory box 2 figure 3) [29]. Dynastic effects could inflate the association between the inherited genotype and phenotype in offspring by capturing social or environmental influences of the parental phenotype. For example, parents with higher educational attainment may be better placed to afford private tuition for their children or buy housing within the catchment area for better performing schools, which will likely influence their children’s educational attainment beyond the effect of inherited genotype. Mendelian randomisation estimates in samples of unrelated individuals may incorrectly attribute the indirect effects of parental phenotype to the offspring’s exposure, biasing the direct association with the outcome of interest [28].

**Explanatory box 2 figure 3.**
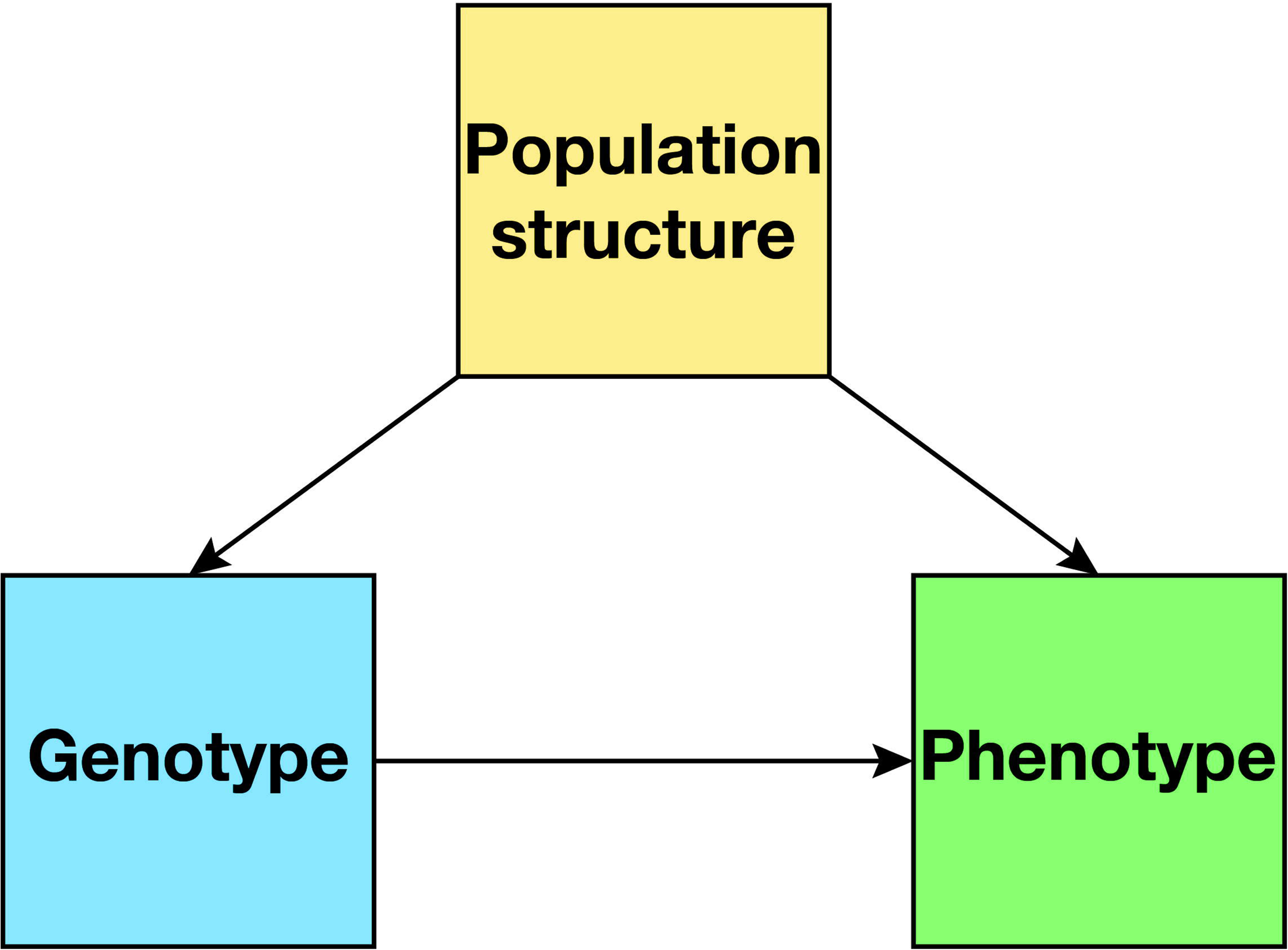
Dynastic effects.

### Within-family Mendelian randomisation

Standard MR assumes that alleles are randomly distributed at the population level. However, this is an approximation and the allocation of alleles is only truly random at the family level, when the alleles are inherited from parents to offspring at conception. Within-family MR therefore uses the family as the unit of study to model or avoid bias due to the non-random distribution of alleles at the population level. Additionally, standard MR assumes that there are no dynastic effects from parent to offspring, while at the family level such effects are assumed to be equal and can therefore be modelled or avoided using within-family designs. Within-family MR designs therefore either model the parental genotypes (such as in parent– offspring designs) or use the random difference between siblings (within-sibship designs) to test direct causal effects [18].

## METHODS

### HUNT

#### Overview

The Trøndelag Health Study (HUNT) is a longitudinal population health survey of adults in the Trøndelag region of Norway and is considered generally representative of the Norwegian population [30]. From commencement, four surveys have been conducted: HUNT1 (1984–86), HUNT2 (1995–97), HUNT3 (2006–08), and HUNT4 (2017–19). Data collected in HUNT includes questionnaires, clinical measurements, and biological samples [31]. We used information on 28 448 siblings from 11 347 sibships who had complete data on genotype, educational attainment, and outcome variables in HUNT2. Siblings were identified using KING software [32], with pairs defined as follows: kinship coefficient between 0.177 and 0.355; the proportion of the genomes that share two alleles identity by descent (IBD) >0.08; and the proportion of the genome that share zero alleles IBD >0.04 [18].

#### Phenotype data

Information regarding educational attainment was obtained from the HUNT2 questionnaire. Participants answered the question, ‘What is your highest level of education?’ by selecting one of five categories: i) ‘Primary school’, ii) ‘High school or equivalent for 1–2 years’, iii) ‘Completed high school or equivalent’, iv) ‘University or equivalent for <4 years’, v) ‘University or college for ≥4 years’. We aligned the HUNT education categories with the International Standard Classification of Education (ISCED) [33] and converted to years of schooling. Individuals who completed primary school were assigned 10 years, those who attended high school for 1–2 years were assigned 12 years, those who completed high school 13 years, those with university degrees 16 years.

Serum concentrations of total cholesterol (TC), high-density lipoprotein cholesterol (HDL-C), and triglycerides (TG) were obtained from non-fasted samples of fresh blood.We calculated low-density lipoprotein cholesterol (LDL-C) concentration using the Friedewald equation (TC – HDL-C – [TG / 2.2]) [34] with lipid values obtained in the same HUNT survey and having excluded participants with triglycerides concentrations >4.5 mmol·L^−1^ [35]. For participants who had data on educational attainment but were missing lipid values measured in HUNT2, we used lipid measurements from HUNT3 if available.

Information regarding diagnosed CVD events (main, not secondary, diagnoses) were obtained from electronic patient records of hospital admissions at Levanger Hospital, Namsos Hospital, and St Olav’s University Hospital in the Nord-Trøndelag Hospital Trust region. A CVD event was defined according to the International Classification of Diseases (ICD-9 codes 410, 430–434, 436; ICD-10 codes I21, I22, I60–I64, excluding I63.6).

Genotyping

There were 70 517 genotyped individuals who participated in either HUNT2 or HUNT3 [36]. Standard HumanCoreExome arrays (HumanCoreExome12 v1.0 and v1.1) were used to genotype 12 864 samples and a customised HumanCoreExome array (UM HUNT Biobank v1.0) for the remainder. Quality control was performed separately for genotype data from different arrays. The call rate of genotyped samples was >99%. Imputation was performed on samples of recent European ancestry using Minimac3 (v2.0.1) [37] from a merged reference panel constructed from the Haplotype Reference Consortium panel (release version 1.1) [38] and a merged local reference panel based on 2201 whole-genome sequenced HUNT participants [39].

### UK Biobank

Overview

UK Biobank is a prospective population study of over 500 000 individuals aged between 40–69 years old at the time of baseline data collection (2006–10) [40]. Participants answered questionnaires, underwent clinical measurements and imaging, and provided biological samples. We used data on 27 040 siblings from 13 180 sibships who had complete data on genotype, educational attainment, and outcome variables. Siblings pairs were identified as per a previous study using UK Biobank-derived estimated of pairwise identical by state (IBS) kinships and the proportion of unshared loci (IBS0) [18,41]. We defined sibling pairs according to bounds for IBS (0.5–21 * IBS0 < IBS < 0.7) and IBS0 (0.001 < IBS0 < 0.008) [42].

### Phenotype data

Participants in UK Biobank answered a touchscreen questionnaire. Information on educational attainment was captured with the question, ‘Which of the following qualifications do you have?’ by selecting one or more of: i) ‘College or university degree’, ii) ‘A levels/AS levels or equivalent’, iii) ‘O levels/GCSEs or equivalent’, iv) ‘CSEs or equivalent’, v) ‘NVQ or HND or HNC or equivalent’, vi) ‘Other professional qualifications eg: nursing, teaching’, vii) ‘None of the above’. We assigned years of education as per a previous publication [19] that used the ISCED classification. Individuals who obtained a college or university degree were assigned 20 years, those who completed A levels/AS levels or equivalent 13 years, those who completed O levels/GCSEs or equivalent 10 years, those who completed CSEs or equivalent 10 years, NVQ/HND/HNC or equivalent 19 years, those with other professional qualifications 15 years, and 7 years for those who answered ‘none of the above’. Participants who selected ‘Prefer not to answer’ were excluded.

Serum concentrations of HDL-C, direct LDL-C, and TG were obtained from non-fasted serum blood samples [43].

Diagnosed CVD events were identified from data on hospital inpatient admissions, obtained through linkage to external data providers. We included only main, not secondary, diagnoses in our analysis and defined a CVD event using the same ICD-9 and ICD-10 codes as with the HUNT cohort.

### Genotyping

There were 488 377 genotyped individuals in UK Biobank [44]. Genotyping was performed using UK BiLEVE (*n* = 49 950) and UK Biobank (*n* = 438 427) Axiom arrays. SHAPEIT3 software was used for pre-phasing [45]. Imputation was performed using Impute4 software and Haplotype Reference Consortium [38] and UK10K [46] reference panel.

### Polygenic score

The educational attainment polygenic score (PGS) is a continuous liability to years spent in formal education. We created a weighted allele score of single nucleotide polymorphisms (SNPs) identified as significant at the genome-wide level (*P* < 5×10^−8^) in a recent GWAS of educational attainment [47]. HUNT participants were not included in this meta-analysis. The summary data was LD clumped (*r*^2^ < 0.001, physical distance threshold = 10 000 kb) using PLINK2 [48]. We removed SNPs with a mean allele frequency (MAF) ≤ 0.01 and *R*^2^ ≤ 0.80 (HUNT) or INFO ≤ 0.80 (UK Biobank) to obtain 592 independent genetic variants present in both the HUNT and UK Biobank cohorts. Each participant’s PGS is their allelic dosage multiplied by the estimated coefficient for each genetic variant from Okbay et al. [47], summed across all genetic variants [49]. We calculated the *F* statistic for the regression of educational attainment on the PGS to assess the risk of weak instrument bias.

### Statistical analysis

#### Individual-level analysis

For each individual-level analysis, the HUNT and UK Biobank cohorts were modelled separately then meta-analysed using a fixed-effects model. We weighted by sample size using the estimates and standard errors from the fitted models of the individual cohorts.

##### Population and within-sibship models

The population model was a regression of the individual’s measured outcome on the exposure. For the within-sibship models, we centred each sibling’s exposure by subtracting the mean sibship exposure value from the individual’s scaled exposure value and included this sibship mean exposure in the population model as a fixed effect to account for variation between siblings [18,28]. All measurements were scaled to have a standard deviation (SD) of Using the within-sibship estimator as fixed effect has previously been used in Cox proportional hazards regression [42,50,51]. To account for collinearity between siblings, we calculated robust standard errors clustered on the family identifier using a sandwich estimator for both population and within-sibship models.

##### Phenotype models

We regressed serum lipid concentrations and incident CVD against measured educational attainment using linear regression and Cox regression, respectively. We treated educational attainment as a continuous measure scaled to SD units and *z*-standardised (mean = 0, SD = 1) the lipid variables. We included age and sex as covariates. In the within-sibship model, we centred educational attainment (scaled to SD units) by subtracting the mean sibship value (also scaled to SD units), and included the mean sibship education as a fixed effect in addition to age and sex.

For the Cox regression, we used date of participation in HUNT2 or UK Biobank as the baseline. For participants who had one or more recorded CVD events, the follow-up interval was calculated to the first diagnosed episode subsequent to participation in either cohort. For participants without a recorded CVD event, the follow-up interval was calculated to date of death or the most recent date of follow-up in their respective cohort (HUNT: 01.12.2019, UK Biobank: 31.03.2021). We retained individuals who had recorded a CVD event prior to their participation in either cohort, though they were treated as if they had not received a diagnosis and their interval calculated to their first event after participation.

##### PGS-phenotype models

We estimated the associations of a PGS of educational attainment on measured educational attainment and all outcomes. We scaled the PGS to SD units for analysis. Measured educational attainment was scaled to SD units and the lipid variables *z*-standardised. In addition to age, sex, and genotyping batch, we included the first 20 genetic principal components to adjust for population structure. In the within-sibship model, we centred the PGS (scaled to SD units) by subtracting the mean sibship value (also scaled to SD), and included the mean sibship PGS as a fixed effect in addition to the abovementioned covariates.

##### 1-sample Mendelian randomisation models

First, we estimated the PGS-education association by regressing measured educational attainment on the educational attainment PGS, both scaled to SD units. We then regressed each outcome on the fitted values from the PGS-education model, using linear regression for the lipid measures and Cox regression for CVD. Each regression analysis was adjusted for age, sex, genotyping batch, and the first 20 genetic principal components. The within-sibship models were additionally adjusted for the within-sibship mean for the PGS. We used two-stage least squares to calculate the point estimate.

#### Summary-level analysis

##### 2-sample Mendelian randomisation models

We used summary data from a recent within-sibship GWAS meta-analysis of phenotypes in 19 cohorts, including HUNT and UK Biobank [20]. The participating cohorts were from Europe (*n* = 12), North America (*n* = 4), Australia (*n* = 2), and China (*n* = 1), and predominantly of European ancestry. For the GWAS analyses, data was available for 178 076 individuals from 77 832 sibships. In our analysis, we used data from the European ancestry cohort meta-analysis for education (*n* = 128 777), LDL-C (*n* = 82 044), HDL-C (*n* = 97 266), and TG (*n* = 92 587). For each cohort, the lipid variables were *z*-standardised prior to GWAS analysis. In the GWAS meta-analyses, years in full-time education the lipid measures were scaled to SD units. For the CVD analysis, the within-sibship summary statistics were obtained from the HUNT population only (*n* = 38 577) and logistic regression, as opposed to Cox regression, was used. There is near-complete sample overlap given that the within-sibship GWASs were conducted in the same cohorts. As our instrument, we used the same 592 genetic variants as in the PGS-outcome and 1-sample MR analyses and obtained effect estimates using an inverse variance weighted (IVW) approach.

All analyses were performed in R version 4.1.3 [52]. The packages data.table (1.14.6), dplyr (1.0.10), foreign (0.8-83), lubdridate (1.8.0), magrittr (2.0.3), purrr (0.3.5), stringr (1.5.0), tibble (3.1.8), and tidyr (1.2.1) were used for data import and wrangling; AER (1.2-10), broom (1.0.1), lmtest (0.9-40), metafor (3.8-1), sandwich (3.0-2), survival (3.4-0), and TwoSampleMR (0.5.6) for analysis; and ggplot2 (3.4.0) and patchwork (1.1.2) for plot creation and design.

## RESULTS

Descriptive characteristics of the two cohorts are given in Table 1.

**Table 1.** Characteristics of participants from HUNT and UK Biobank.

|  | **HUNT** |  | **UK Biobank** |  |
| --- | --- | --- | --- | --- |
|  | **N (%)** | **Mean (SD)** | **N (%)** | **Mean (SD)** |
| Participants | 28 448 |  | 27 040 |  |
| Sibships | 11 347 |  | 13 180 |  |
| Female | 14 678 (51.6) |  | 15 493 (57.3) |  |
| Age |  | 48.5 (15.4) |  | 57.1 (7.4) |
| Education (years) |  | 12.0 (2.1) |  | 13.5 (5.1) |
| LDL-C (mmol·L<sup>-1</sup>) |  | 3.7 (1.1) |  | 3.6 (0.9) |
| HDL-C (mmol·L<sup>-1</sup>) |  | 1.4 (0.4) |  | 1.5 (0.4) |
| Triglycerides (mmol·L<sup>-1</sup>) |  | 1.7 (1.0) |  | 1.7 (1.0) |
| CVD cases | 4148 (14.6) |  | 1329 (4.9) |  |
| Follow-up interval (days) |  | 7342 (2271) |  | 4318 (788) |

CVD = cardiovascular disease, HDL-C = high-density lipoprotein cholesterol, LDL-C = low- density lipoprotein cholesterol, N = number, SD = standard deviation.

### Reported educational attainment, lipids, and cardiovascular disease

We estimated the associations of educational attainment with serum lipid concentrations and CVD risk using population and within-sibship models (Figures 1–4 and Supplementary Tables S1–4). In the within-sibship models, a 1 SD (2.1 years in HUNT, 5.1 years in UK Biobank) higher attained education was associated with lower concentrations of LDL-C (–0.03 SD; 95% CI –0.04 to –0.02) and TG (–0.04 SD; 95% CI –0.05 to –0.03), higher concentrations of HDL-C (0.01 SD; 95% CI 0.00 to 0.02), and a lower risk of CVD (HR 0.94; 95% CI 0.89 to 1.00).

**Figure 1.**
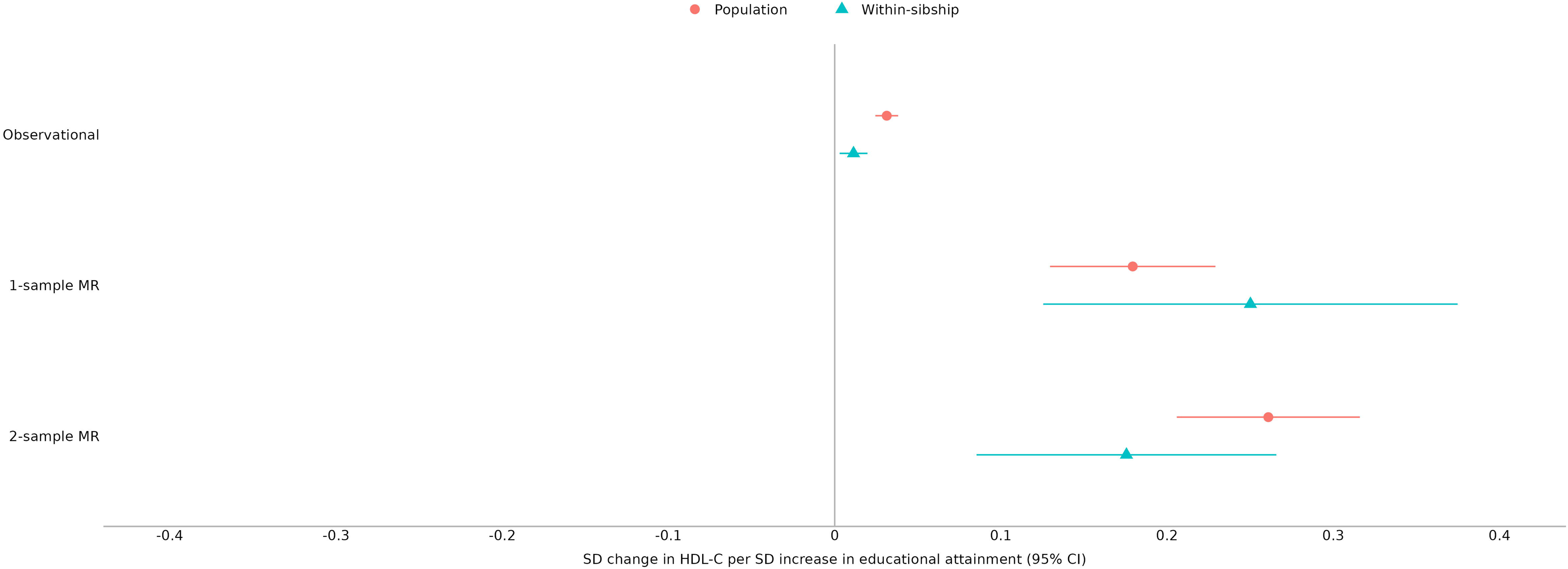
Educational attainment and low-density lipoprotein cholesterol. Estimated associations between educational attainment and low-density lipoprotein cholesterol concentration using both observational and Mendelian randomisation models. The estimates are presented in SD units. Abbreviations: CI = confidence interval, LDL-C = low-density lipoprotein cholesterol, MR = Mendelian randomisation, SD = standard deviation.

### Individual participant Mendelian randomisation

The *F* statistic for the educational attainment PGS was 418.0, suggesting low risk of weak instrument bias, and the *R^2^* = 1.4%. The educational attainment PGS was strongly associated with measured educational attainment in population (0.15 SD; 95% CI 0.14 to 0.16) and within-sibship models (0.08 SD; 95% CI 0.06 to 0.09) (Figure 5 and Supplementary Table S5). In population models, the educational attainment PGS was associated with lower serum concentrations of LDL-C (–0.02 SD; 95% CI –0.03 to –0.01) and TG (–0.04 SD; 95% CI –0.05 to –0.03), higher serum concentrations of HDL-C (0.03 SD; 95% CI 0.02 to 0.03), and a lower risk of CVD (HR 0.94; 95% CI 0.91 to 0.97). In the within-sibship models, the association estimates for serum concentrations of TG (–0.03 SD; 95% CI –0.04 to –0.02) and HDL-C (0.02 SD; 95% CI 0.01 to 0.03) were consistent with those of the population models. As expected, the estimates were less precise due to lower statistical power in the sibling models. There was little evidence from the within-sibship estimates that the scores associated with serum LDL-C concentration (0.00 SD; 95% CI – 0.01 to 0.01). Higher educational attainment PGS was associated with a lower risk of CVD within sibships (HR 0.93; 95% CI 0.88 to 0.99).

**Figure 2.**
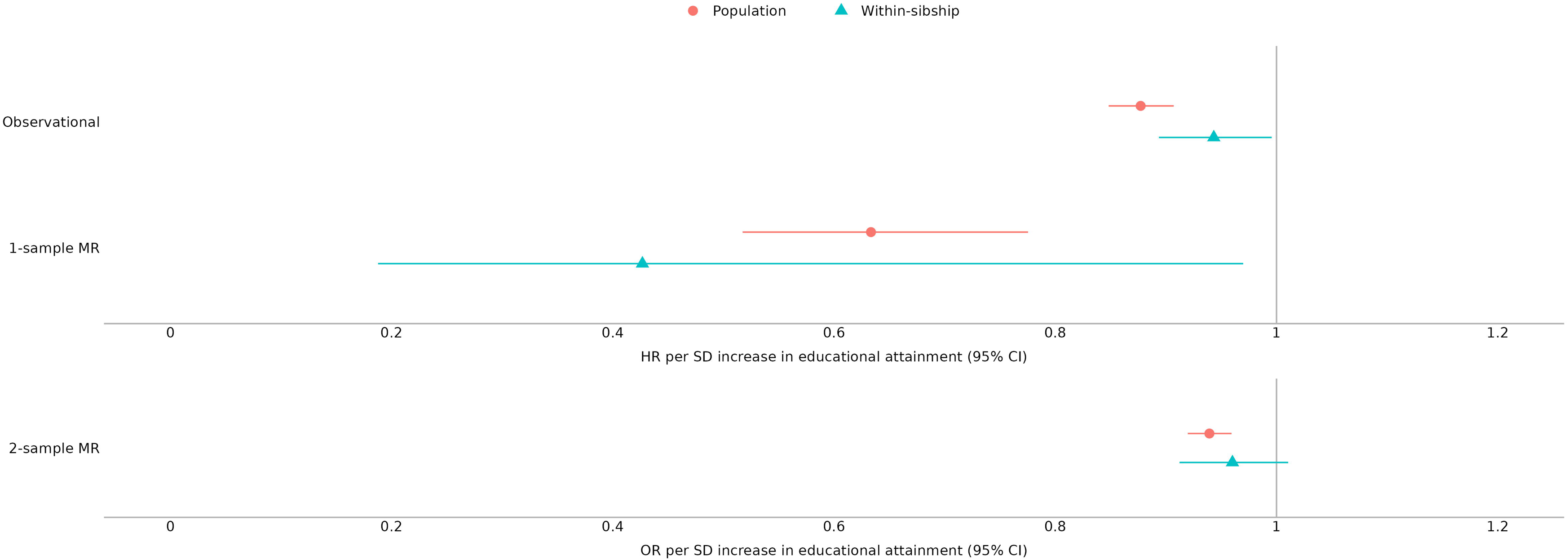
Educational attainment and high-density lipoprotein cholesterol. Estimated associations between educational attainment and high-density lipoprotein cholesterol concentration using both observational and Mendelian randomisation models. The estimates are presented in SD units. Abbreviations: CI = confidence interval, HDL-C = high-density lipoprotein cholesterol, MR = Mendelian randomisation, SD = standard deviation.

We subsequently estimated the causal effects of educational attainment on each outcome (Figures 1–4 and Supplementary Tables S1–4). A 1 SD increase in educational attainment was associated with decreased serum levels of LDL-C (–0.12 SD, 95% CI –0.18 to –0.07) and TG (–0.26 SD, 95% CI –0.32 to –0.20), increased levels of HDL-C (0.18 SD, 95% CI 0.13 to 0.23), and lowered the risk of CVD (HR 0.63, 95% CI 0.52 to 0.78). In the within-sibship models, the effect estimates for TG (–0.39 SD, 95% CI –0.59 to –0.20), HDL-C (0.25 SD, 95% CI 0.13 to 0.37) and CVD risk (HR 0.43, 95% CI 0.19 to 0.97) were broadly consistent with those of the population models but imprecise. The estimate for LDL-C (–0.02 SD, 95% CI –0.19 to 0.15) was attenuated towards the null, though not completely and consistent in direction with the phenotypic associations.

**Figure 3.**
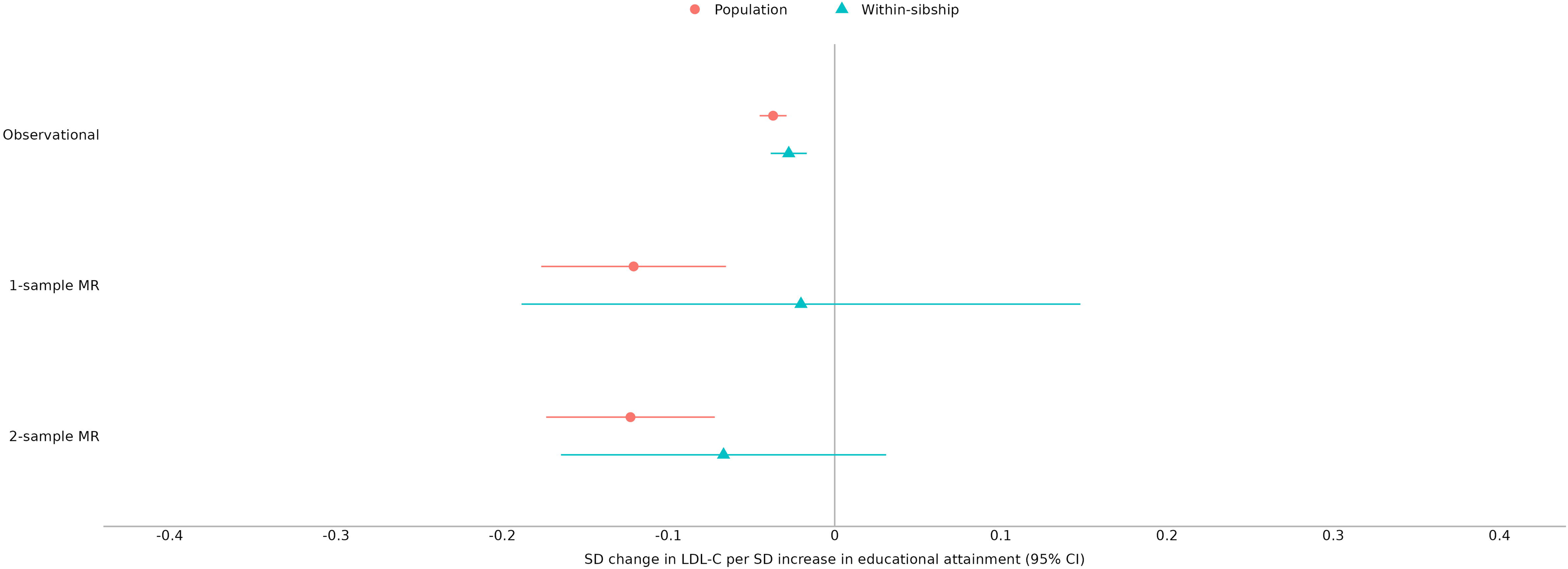
Educational attainment and triglycerides. Estimated associations between educational attainment and triglycerides concentration using both observational and Mendelian randomisation models. The estimates are presented in SD units. Abbreviations: CI = confidence interval, MR = Mendelian randomisation, SD = standard deviation, TG = triglycerides.

### Mendelian randomisation using summary data

We performed 2-sample MR analyses of educational attainment and outcomes using summary data from a recent within-sibship GWAS meta-analysis (Figures 1–4 and Supplementary Tables S1–4) [20]. In the population models, higher levels of educational attainment decreased serum LDL-C (–0.12 SD, 95% CI –0.17 to –0.07) and TG (–0.26 SD, 95% CI –0.32 to –0.21) concentrations, increased serum HDL-C (0.26 SD, 95% CI 0.21 to 0.32) concentrations and were protective against CVD risk (OR 0.94, 95% CI 0.92 to 0.96). For each outcome, estimates were smaller for the within-sibship models compared to the respective population models.

**Figure 4.**
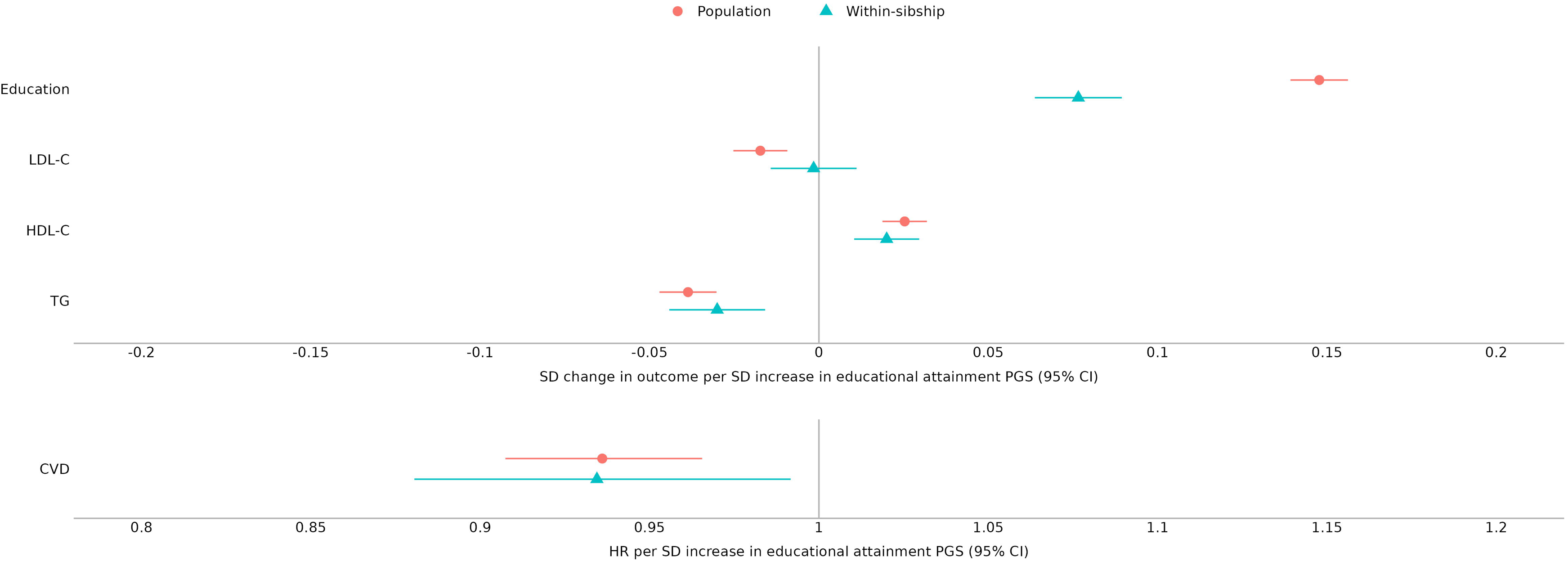
Educational attainment and cardiovascular disease. Estimated associations between educational attainment and cardiovascular disease using both observational and Mendelian randomisation models. The estimates are presented as hazard ratios in the individual-level models and odds ratios in the summary-level models. Abbreviations: CI = confidence interval, HR = hazard ratio, MR = Mendelian randomisation, OR = odds ratio, SD = standard deviation.

**Figure 5.**
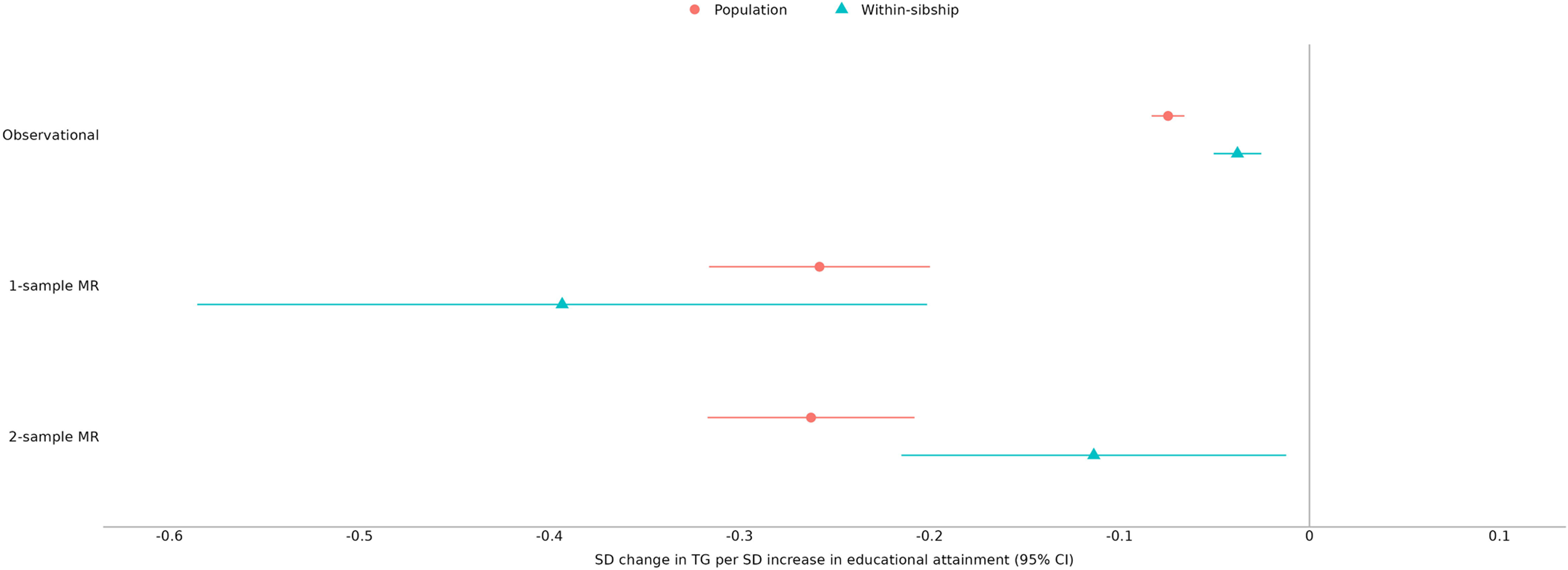
Educational attainment polygenic score and five outcomes. Estimated associations between an educational attainment polygenic score and five outcomes for population and within-sibship models. The estimates for cardiovascular disease are presented as hazard ratios and the remaining outcomes as SD units. Abbreviations: CI = confidence interval, CVD = cardiovascular disease, HDL-C = high-density lipoprotein cholesterol, HR = hazard ratio, LDL-C = low-density lipoprotein cholesterol, PGS = polygenic score, SD = standard deviation, TG = triglycerides.

## DISCUSSION

Using sibling data from HUNT, UK Biobank, and a large within-sibship GWAS meta-analysis, we examined the effects of educational attainment on serum lipid concentrations and CVD risk. Across our analyses, we found higher educational attainment to be beneficial for HDL-C and TG concentrations in adulthood. We also found evidence that higher educational attainment reduced CVD risk. Our within-sibship estimate of the effect of educational attainment on LDL-C were imprecise but comparable to the population models suggesting a causal reduction. Generally, there was a trend for attenuation, albeit small, in within-sibship compared to population models, which could indicate that the causal effect of educational attainment is slightly weaker than previously reported.

### Comparison with other studies

The results of within-sibship models are consistent with previous studies that used instrumental variable approaches to analyse population data. Two MR analyses using summary-level data demonstrated a protective effect of educational attainment on CHD [12,13], ischaemic stroke [13], and beneficial effects on HDL-C and TG concentrations [12]. Similarly, a quasi-experimental analysis, which used differences in state-level compulsory schooling laws (CSLs) in the United States as instrumental variables for educational attainment, found higher levels were protective for self-reported heart disease and beneficial for HDL-C and TG concentrations [53]. As in our results, the associations of educational attainment with HDL-C and TG were far stronger in the instrumental variable analysis compared to the observational analysis and suggests that observational models may underestimate the effects on these lipid measures [53]. However, in the two studies that analysed lipids there were no apparent benefits to LDL-C concentration [12,53], whereas our results suggest some benefit.

In our analyses, any attenuation of the population estimate in the corresponding within-sibship analysis is generally small and suggests weak to little evidence of substantial biases from demographic or dynastic effects. This is consistent with another study using within-sibship MR to examine the effects of educational attainment on all-cause mortality and other health outcomes [42], and one of height on CHD and cancer risk [54]. Other recent studies examining the effects of height and BMI on educational attainment have demonstrated marked attenuation of estimates in within-sibship models, indicating that the comparative population models were likely biased by demographic or dynastic effects and make a strong case for using within-sibship MR as a sensitivity test where the assumptions of MR are unlikely to hold in unrelated individuals [18,20].

Similar to our findings, reduced but not completely attenuated estimates have been demonstrated in observational studies using sibling designs. In a study of over 800 000 Norwegian siblings [55], a one-category decrease in level of education was associated with an increased premature CVD mortality hazard rate in men and women. In the sibling analysis, the hazard rates were lower but not completely attenuated, suggesting that at least some of the educational inequalities are a consequence of factors shared by siblings. Another sibling comparison study of CVD risk factors, including total serum cholesterol concentration, reported that approximately one third of the educational gradient for each outcome was explained by factors shared between siblings [56].

Despite substantial secular rises in educational attainment in certain countries and corresponding declines in population prevalence of certain CVD risk factors [57], differences in prevalence between educational groups contribute to persistent inequalities in CVD risk [58–61]. For example, a study of three Norwegian birth cohorts showed that two thirds of the excess premature CVD mortality in the lowest compared to highest education group were mediated by unfavourable CVD risk factors for all analysed birth cohort groups [62].

Interestingly, BMI was not found to be an important mediator in any of the cohort groups, potentially due to the generally small differences between education groups and healthy BMIs across cohorts. Another mediation analysis using complimentary observational and multivariable MR approaches in UK Biobank participants showed that three CVD risk factors (smoking, BMI, and systolic blood pressure) individually explain approximately 10–20% of the effect of educational attainment on CVD risk, and combined may explain over 40% [63]. Though the relationship between educational attainment, levels of CVD risk factors and CVD risk are likely driven by several underlying mechanisms, findings across observational, sibling, regression discontinuity and MR study designs provide convincing evidence for a causal effect of educational attainment that cannot solely be explained by familial factors shared by siblings.

### Strengths and limitations of this study

The principal strengths of our work are using the within-sibship MR design in HUNT, UK Biobank, and a large within-sibship GWAS meta-analysis [20], and triangulation of evidence using complimentary modelling approaches: phenotypic, genetic, and within-sibship [64]. There are also a number of limitations. Our individual-level within-sibship estimates were considerably less precise than the population estimates [18]. However, the summary-level analyses did improve the precision of the estimates where data was available [20], and estimates from both approaches were generally consistent with the population estimates. Future studies should aim to increase availability of within-family data in biobanks and conduct sibship GWASs to improve the precision of within-sibship MR estimates and expand the potential for its use as a sensitivity analysis of conventional population estimates. Participants in our data sources were predominantly of European ancestry, which likely limits the generalisability of our results to other populations and different social contexts. The genetic variants included in our PGS are likely to influence other domains of educational attainment rather than just years spent in education, hence the estimates are unlikely to reflect purely the effects of an additional year of education [21]. There was a high degree of overlap between the exposure and outcome samples in our summary-level analyses, which would bias the MR estimates towards the observational results. However, sample overlap bias with SNPs that are strongly associated with the exposure (education) is generally small. There is evidence for selection on education in biobanks which could induce associations with our study outcomes if they are also related to biobank participation [65–67]. Lastly, although less affected by the confounds of population GWASs, family-based GWASs can suffer from certain sources of confounding, such as indirect genetic effects from siblings [68,69].

## CONCLUSION

Our results suggest that previously reported beneficial effects of educational attainment on serum lipid concentrations and CVD risk are likely causal and not due to bias from demographic or dynastic effects. Overall, this study suggests that increasing education may be beneficial for cardiovascular health.

## Data Availability

All data produced in the present study are available upon reasonable request to the authors.

https://hunt-db.medisin.ntnu.no/hunt-db/

https://gwas.mrcieu.ac.uk/

## DECLARATIONS

### Ethics approval

The use of HUNT data was approved by the Mid-Norway Regional Committee for Medical and Health Research Ethics (2015/1197) and South East Norway Regional Committee for Medical and Health Research Ethics (2017/2479). All participants signed informed consent for participation and the use of data in research. UK Biobank obtained ethics approval from the North West Multi-centre Research Ethics Committee and obtained informed consent from all study participants.

### Data availability

The HUNT data reported in this study cannot be deposited in a public repository because it is governed by Norwegian law. To request access, researchers associated with Norwegian research institutes can apply for the use of HUNT data and samples with approval by the Regional Committee for Medical and Health Research Ethics. Researchers from other countries may apply if collaborating with a Norwegian Principal Investigator. Information for data access can be found at https://www.ntnu.edu/hunt/data. The HUNT variables are available for browsing on the HUNT databank at https://hunt-db.medisin.ntnu.no/hunt-db/. Use of the full genetic dataset requires the use of an approved secure computing solution such as the HUNT Cloud (https://docs.hdc.ntnu.no). UK Biobank individual-level participant data are available via enquiry to. Summary data from the within-sibship meta-analysis GWAS are publicly available for download on OpenGWAS (https://gwas.mrcieu.ac.uk/) via the **TwoSampleMR** R package. Note that the summary data include both ‘population’ and ‘within-sibship’ estimates for each phenotype, with the model detailed in the metadata notes.

### Availability of data and materials

Supplementary data are available online.

### Author contributions

BMB and ØEN conceived the study. BMB, PRJ and ØEN designed the study. BMB and ØEN obtained funding. BMB, PRJ and ØEN wrote the analysis plan. PRJ performed the analysis. LB performed the UK Biobank analysis. PRJ drafted the original manuscript. All co-authors contributed to interpretation of results, refined the manuscript, and approved its final version. BMB and ØEN are joint senior authors. The corresponding author attests that all listed authors meet authorship criteria and that no others meeting the criteria have been omitted.

## Acknowledgements

The Trøndelag Health Study (HUNT) is a collaboration between HUNT Research Centre (Faculty of Medicine and Health Sciences, NTNU, Norwegian University of Science and Technology), Trøndelag County Council, Central Norway Regional Health Authority and the Norwegian Institute of Public Health. The genotyping in HUNT was financed by the National Institutes of Health; University of Michigan; the Research Council of Norway; the Liaison Committee for Education, Research and Innovation in Central Norway; and the Joint Research Committee between St Olavs hospital and the Faculty of Medicine and Health Sciences, NTNU. The K.G. Jebsen Center for Genetic Epidemiology is financed by Stiftelsen Kristian Gerhard Jebsen; Faculty of Medicine and Health Sciences, NTNU, Norway. This research has been conducted using the UK Biobank Resource under application number 40135. LB received support from the K.G. Jebsen Center for Genetic Epidemiology funded by Stiftelsen Kristian Gerhard Jebsen; Faculty of Medicine and Health Sciences, NTNU; The Liaison Committee for education, research and innovation in Central Norway; and the Joint Research Committee between St Olavs Hospital and the Faculty of Medicine and Health Sciences, NTNU.

## REFERENCES

[1] Timmis A, Vardas P, Townsend N, Torbica A, Katus H, De Smedt D, et al. European Society of Cardiology: cardiovascular disease statistics 2021. European Heart Journal 2022;43:716–99. 10.1093/eurheartj/ehab892.

[2] Veronesi G, Ferrario MM, Kuulasmaa K, Bobak M, Chambless LE, Salomaa V, et al. Educational class inequalities in the incidence of coronary heart disease in Europe. Heart 2016;102:958. 10.1136/heartjnl-2015-308909.

[3] Kubota Y, Heiss G, MacLehose RF, Roetker NS, Folsom AR. Association of Educational Attainment With Lifetime Risk of Cardiovascular Disease: The Atherosclerosis Risk in Communities Study. JAMA Internal Medicine 2017;177:1165–72. 10.1001/jamainternmed.2017.1877.

[4] Pierce JP, Fiore MC, Novotny TE, Hatziandreu EJ, Davis RM. Trends in Cigarette Smoking in the United States: Educational Differences Are Increasing. JAMA 1989;261:56–60. 10.1001/jama.1989.03420010066034.

[5] Latvala A, Rose RJ, Pulkkinen L, Dick DM, Korhonen T, Kaprio J. Drinking, smoking, and educational achievement: Cross-lagged associations from adolescence to adulthood. Drug and Alcohol Dependence 2014;137:106–13. 10.1016/j.drugalcdep.2014.01.016.

[6] Lawlor DA, Clark H, Davey Smith G, Leon DA. Childhood intelligence, educational attainment and adult body mass index: findings from a prospective cohort and within sibling-pairs analysis. International Journal of Obesity 2006;30:1758–65. 10.1038/sj.ijo.0803330.

[7] Cohen AK, Rai M, Rehkopf DH, Abrams B. Educational attainment and obesity: a systematic review. Obesity Reviews 2013;14:989–1005. 10.1111/obr.12062.

[8] Hughes A, Wade KH, Dickson M, Rice F, Davies A, Davies NM, et al. Common health conditions in childhood and adolescence, school absence, and educational attainment: Mendelian randomization study. Npj Science of Learning 2021;6:1. 10.1038/s41539-020-00080-6.

[9] Jacobs DR, Woo JG, Sinaiko AR, Daniels SR, Ikonen J, Juonala M, et al. Childhood Cardiovascular Risk Factors and Adult Cardiovascular Events. N Engl J Med 2022;386:1877–88. 10.1056/NEJMoa2109191.

[10] Davey Smith G, Ebrahim S. ‘Mendelian randomization’: can genetic epidemiology contribute to understanding environmental determinants of disease? International Journal of Epidemiology 2003;32:1–22. 10.1093/ije/dyg070.

[11] Lawlor DA, Harbord RM, Sterne JAC, Timpson N, Davey Smith G. Mendelian randomization: Using genes as instruments for making causal inferences in epidemiology. Statistics in Medicine 2008;27:1133–63. 10.1002/sim.3034.

[12] Tillmann T, Vaucher J, Okbay A, Pikhart H, Peasey A, Kubinova R, et al. Education and coronary heart disease: mendelian randomisation study. BMJ 2017;358:j3542. 10.1136/bmj.j3542.

[13] Gill D, Efstathiadou A, Cawood K, Tzoulaki I, Dehghan A. Education protects against coronary heart disease and stroke independently of cognitive function: evidence from Mendelian randomization. International Journal of Epidemiology 2019;48:1468–77. 10.1093/ije/dyz200.

[14] Sanderson E, Davey Smith G, Bowden J, Munafò MR. Mendelian randomisation analysis of the effect of educational attainment and cognitive ability on smoking behaviour. Nature Communications 2019;10:2949. 10.1038/s41467-019-10679-y.

[15] Böckerman P, Viinikainen J, Pulkki-Råback L, Hakulinen C, Pitkänen N, Lehtimäki T, et al. Does higher education protect against obesity? Evidence using Mendelian randomization. Preventive Medicine 2017;101:195–8. 10.1016/j.ypmed.2017.06.015.

[16] Kong A, Thorleifsson G, Frigge ML, Vilhjalmsson BJ, Young AI, Thorgeirsson TE, et al. The nature of nurture: Effects of parental genotypes. Science 2018;359:424. 10.1126/science.aan6877.

[17] Davies NM, Howe LJ, Brumpton B, Havdahl A, Evans DM, Davey Smith G. Within family Mendelian randomization studies. Human Molecular Genetics 2019;28:R170–9. 10.1093/hmg/ddz204.

[18] Brumpton B, Sanderson E, Heilbron K, Hartwig FP, Harrison S, Vie GÅ, et al. Avoiding dynastic, assortative mating, and population stratification biases in Mendelian randomization through within-family analyses. Nature Communications 2020;11:3519. 10.1038/s41467-020-17117-4.

[19] Lee JJ, Wedow R, Okbay A, Kong E, Maghzian O, Zacher M, et al. Gene discovery and polygenic prediction from a genome-wide association study of educational attainment in 1.1 million individuals. Nature Genetics 2018;50:1112–21. 10.1038/s41588-018-0147-3.

[20] Howe LJ, Nivard MG, Morris TT, Hansen AF, Rasheed H, Cho Y, et al. Within-sibship genome-wide association analyses decrease bias in estimates of direct genetic effects. Nature Genetics 2022;54:581–92. 10.1038/s41588-022-01062-7.

[21] Howe LJ, Tudball M, Davey Smith G, Davies NM. Interpreting Mendelian-randomization estimates of the effects of categorical exposures such as disease status and educational attainment. International Journal of Epidemiology 2022;51:948–57. 10.1093/ije/dyab208.

[22] Tudball MJ, Bowden J, Hughes RA, Ly A, Munafò MR, Tilling K, et al. Mendelian randomisation with coarsened exposures. Genetic Epidemiology 2021;45:338–50. 10.1002/gepi.22376.

[23] Davey Smith G, Hemani G. Mendelian randomization: genetic anchors for causal inference in epidemiological studies. Human Molecular Genetics 2014;23:R89–98. 10.1093/hmg/ddu328.

[24] Haycock PC, Burgess S, Wade KH, Bowden J, Relton C, Davey Smith G. Best (but oft-forgotten) practices: the design, analysis, and interpretation of Mendelian randomization studies. The American Journal of Clinical Nutrition 2016;103:965–78. 10.3945/ajcn.115.118216.

[25] Cardon LR, Palmer LJ. Population stratification and spurious allelic association. The Lancet 2003;361:598–604. 10.1016/S0140-6736(03)12520-2.

[26] Haworth S, Mitchell R, Corbin L, Wade KH, Dudding T, Budu-Aggrey A, et al. Apparent latent structure within the UK Biobank sample has implications for epidemiological analysis. Nature Communications 2019;10:333. 10.1038/s41467-018-08219-1.

[27] Hartwig FP, Davies NM, Davey Smith G. Bias in Mendelian randomization due to assortative mating. Genetic Epidemiology 2018;42:608–20. 10.1002/gepi.22138.

[28] Hwang L-D, Davies NM, Warrington NM, Evans DM. Integrating Family-Based and Mendelian Randomization Designs. Cold Spring Harbor Perspectives in Medicine 2021;11:a039503. 10.1101/cshperspect.a039503.

[29] Morris TT, Davies NM, Hemani G, Smith GD. Population phenomena inflate genetic associations of complex social traits. Sci Adv 2020;6:eaay0328. 10.1126/sciadv.aay0328.

[30] Holmen m.fl J. The Nord-Trøndelag Health Study 1995-97 (HUNT 2). Nor J Epidemiol 2011;13. 10.5324/nje.v13i1.305.

[31] Krokstad S, Langhammer A, Hveem K, Holmen T, Midthjell K, Stene T, et al. Cohort Profile: The HUNT Study, Norway. International Journal of Epidemiology 2013;42:968–77. 10.1093/ije/dys095.

[32] Manichaikul A, Mychaleckyj JC, Rich SS, Daly K, Sale M, Chen W-M. Robust relationship inference in genome-wide association studies. Bioinformatics 2010;26:2867–73. 10.1093/bioinformatics/btq559.

[33] UNESCO Institute for Statistics. International Standard Classification of Education: ISCED 2011 2012.

[34] Friedewald WT, Levy RI, Fredrickson DS. Estimation of the Concentration of Low-Density Lipoprotein Cholesterol in Plasma, Without Use of the Preparative Ultracentrifuge. Clinical Chemistry 1972;18:499–502. 10.1093/clinchem/18.6.499.

[35] Mach F, Baigent C, Catapano AL, Koskinas KC, Casula M, Badimon L, et al. 2019 ESC/EAS Guidelines for the management of dyslipidaemias: lipid modification to reduce cardiovascular risk: The Task Force for the management of dyslipidaemias of the European Society of Cardiology (ESC) and European Atherosclerosis Society (EAS). European Heart Journal 2019;41:111–88. 10.1093/eurheartj/ehz455.

[36] Brumpton BM, Graham S, Surakka I, Skogholt AH, Løset M, Fritsche LG, et al. The HUNT study: A population-based cohort for genetic research. Cell Genomics 2022;2:100193. 10.1016/j.xgen.2022.100193.

[37] Das S, Forer L, Schönherr S, Sidore C, Locke AE, Kwong A, et al. Next-generation genotype imputation service and methods. Nat Genet 2016;48:1284–7. 10.1038/ng.3656.

[38] McCarthy S, Das S, Kretzschmar W, Delaneau O, Wood AR, Teumer A, et al. A reference panel of 64,976 haplotypes for genotype imputation. Nat Genet 2016;48:1279–83. 10.1038/ng.3643.

[39] Zhou W, Fritsche LG, Das S, Zhang H, Nielsen JB, Holmen OL, et al. Improving power of association tests using multiple sets of imputed genotypes from distributed reference panels. Genetic Epidemiology 2017;41:744–55. 10.1002/gepi.22067.

[40] Sudlow C, Gallacher J, Allen N, Beral V, Burton P, Danesh J, et al. UK Biobank: An Open Access Resource for Identifying the Causes of a Wide Range of Complex Diseases of Middle and Old Age. PLOS Medicine 2015;12:e1001779. 10.1371/journal.pmed.1001779.

[41] Hill WG, Weir BS. Variation in actual relationship as a consequence of Mendelian sampling and linkage. Genetics Research 2011;93:47–64. 10.1017/S0016672310000480.

[42] Howe LJ, Rasheed H, Jones PR, Boomsma DI, Evans DM, Giannelis A, et al. Educational attainment, health outcomes and mortality: a within-sibship Mendelian randomization study. International Journal of Epidemiology 2023:dyad079. 10.1093/ije/dyad079.

[43] Welsh C, Celis-Morales CA, Brown R, Mackay DF, Lewsey J, Mark PB, et al. Comparison of Conventional Lipoprotein Tests and Apolipoproteins in the Prediction of Cardiovascular Disease. Circulation 2019;140:542–52. 10.1161/CIRCULATIONAHA.119.041149.

[44] Bycroft C, Freeman C, Petkova D, Band G, Elliott LT, Sharp K, et al. The UK Biobank resource with deep phenotyping and genomic data. Nature 2018;562:203–9. 10.1038/s41586-018-0579-z.

[45] O’Connell J, Sharp K, Shrine N, Wain L, Hall I, Tobin M, et al. Haplotype estimation for biobank-scale data sets. Nat Genet 2016;48:817–20. 10.1038/ng.3583.

[46] Walter K, Min JL, Huang J, Crooks L, Memari Y, McCarthy S, et al. The UK10K project identifies rare variants in health and disease. Nature 2015;526:82–90. 10.1038/nature14962.

[47] Okbay A, Wu Y, Wang N, Jayashankar H, Bennett M, Nehzati SM, et al. Polygenic prediction of educational attainment within and between families from genome-wide association analyses in 3 million individuals. Nat Genet 2022;54:437–49. 10.1038/s41588-022-01016-z.

[48] Purcell S, Neale B, Todd-Brown K, Thomas L, Ferreira MAR, Bender D, et al. PLINK: a tool set for whole-genome association and population-based linkage analyses. Am J Hum Genet 2007;81:559–75. 10.1086/519795.

[49] Allegrini AG, Baldwin JR, Barkhuizen W, Pingault J-B. Research Review: A guide to computing and implementing polygenic scores in developmental research. Journal of Child Psychology and Psychiatry 2022;63:1111–24. 10.1111/jcpp.13611.

[50] Finegood ED, Briley DA, Turiano NA, Freedman A, South SC, Krueger RF, et al. Association of Wealth With Longevity in US Adults at Midlife. JAMA Health Forum 2021;2:e211652. 10.1001/jamahealthforum.2021.1652.

[51] Sjölander A, Lichtenstein P, Larsson H, Pawitan Y. Between–within models for survival analysis. Statistics in Medicine 2013;32:3067–76. 10.1002/sim.5767.

[52] R Core Team. R: A language and environment for statistical computing 2020.

[53] Hamad R, Nguyen TT, Bhattacharya J, Glymour MM, Rehkopf DH. Educational attainment and cardiovascular disease in the United States: A quasi-experimental instrumental variables analysis. PLOS Medicine 2019;16:e1002834. 10.1371/journal.pmed.1002834.

[54] Howe LJ, Brumpton B, Rasheed H, Åsvold BO, Davey Smith G, Davies NM. Taller height and risk of coronary heart disease and cancer: A within-sibship Mendelian randomization study. ELife 2022;11:e72984. 10.7554/eLife.72984.

[55] Næss Ø, Hoff DA, Lawlor D, Mortensen LH. Education and adult cause-specific mortality—examining the impact of family factors shared by 871 367 Norwegian siblings. International Journal of Epidemiology 2012;41:1683–91. 10.1093/ije/dys143.

[56] Ariansen I, Mortensen LH, Graff-Iversen S, Stigum H, Kjøllesdal MKR, Næss Ø. The educational gradient in cardiovascular risk factors: impact of shared family factors in 228,346 Norwegian siblings. BMC Public Health 2017;17:281. 10.1186/s12889-017-4123-0.

[57] Harper S, Lynch J, Smith GD. Social Determinants and the Decline of Cardiovascular Diseases: Understanding the Links. Annu Rev Public Health 2011;32:39–69. 10.1146/annurev-publhealth-031210-101234.

[58] Nordahl H, Rod NH, Frederiksen BL, Andersen I, Lange T, Diderichsen F, et al. Education and risk of coronary heart disease: assessment of mediation by behavioral risk factors using the additive hazards model. Eur J Epidemiol 2013;28:149–57. 10.1007/s10654-012-9745-z.

[59] Ernstsen L, Strand BH, Nilsen SM, Espnes GA, Krokstad S. Trends in absolute and relative educational inequalities in four modifiable ischaemic heart disease risk factors: repeated cross-sectional surveys from the Nord-Trøndelag Health Study (HUNT) 1984– 2008. BMC Public Health 2012;12:266. 10.1186/1471-2458-12-266.

[60] Schultz WM, Kelli HM, Lisko JC, Varghese T, Shen J, Sandesara P, et al. Socioeconomic Status and Cardiovascular Outcomes. Circulation 2018;137:2166–>78. 10.1161/CIRCULATIONAHA.117.029652.

[61] Capewell S, Graham H. Will Cardiovascular Disease Prevention Widen Health Inequalities? PLOS Medicine 2010;7:e1000320. 10.1371/journal.pmed.1000320.

[62] Ariansen I, Strand BH, Kjøllesdal MKR, Steingrímsdóttir ÓA, Mortensen LH, Stigum H, et al. The educational gradient in premature cardiovascular mortality: Examining mediation by risk factors in cohorts born in the 1930s, 1940s and 1950s. European Journal of Preventive Cardiology 2019;26:1096–103. 10.1177/2047487319826274.

[63] Carter AR, Gill D, Davies NM, Taylor AE, Tillmann T, Vaucher J, et al. Understanding the consequences of education inequality on cardiovascular disease: mendelian randomisation study. BMJ 2019;365:l1855. 10.1136/bmj.l1855.

[64] Lawlor DA, Tilling K, Davey Smith G. Triangulation in aetiological epidemiology. International Journal of Epidemiology 2016;45:1866–86. 10.1093/ije/dyw314.

[65] Munafò MR, Tilling K, Taylor AE, Evans DM, Davey Smith G. Collider scope: when selection bias can substantially influence observed associations. International Journal of Epidemiology 2018;47:226–35. 10.1093/ije/dyx206.

[66] Tyrrell J, Zheng J, Beaumont R, Hinton K, Richardson TG, Wood AR, et al. Genetic predictors of participation in optional components of UK Biobank. Nat Commun 2021;12:886. 10.1038/s41467-021-21073-y.

[67] Fry A, Littlejohns TJ, Sudlow C, Doherty N, Adamska L, Sprosen T, et al. Comparison of Sociodemographic and Health-Related Characteristics of UK Biobank Participants With Those of the General Population. American Journal of Epidemiology 2017;186:1026–34. 10.1093/aje/kwx246.

[68] Howe LJ, Evans DM, Hemani G, Smith GD, Davies NM. Evaluating indirect genetic effects of siblings using singletons. PLOS Genetics 2022;18:e1010247. 10.1371/journal.pgen.1010247.

[69] Veller C, Coop G. Interpreting population and family-based genome-wide association studies in the presence of confounding 2023:2023.02.26.530052. 10.1101/2023.02.26.530052.

