## Supplementary for "Education and cardiovascular disease: a within-family Mendelian randomisation analysis"

**Table S1. Educational attainment and low-density lipoprotein cholesterol**

| **Model** | | **Change in outcome per SD increase in educational attainment (SD, 95% CI)** | | |
|---|---|---|---|---|
| | | HUNT | UK Biobank | Combined |
| Observational model | Population | –0.08 (–0.09 to –0.07) | 0.01 (–0.01 to 0.02) | –0.04 (–0.05 to –0.03) |
| | Within-sibship | –0.05 (–0.07 to –0.04) | 0.00 (–0.02 to 0.01) | –0.03 (–0.04 to –0.02) |
| 1-sample MR | Population | –0.16 (–0.25 to –0.07) | –0.08 (–0.15 to –0.02) | –0.12 (–0.18 to –0.07) |
| | Within-sibship | –0.02 (–0.23 to 0.18) | –0.02 (–0.29 to 0.25) | –0.02 (–0.19 to 0.15) |
| | | | | Consortium |
| 2-sample MR | Population | | | –0.12 (–0.17 to –0.07) |
| | Within-sibship | | | –0.07 (–0.16 to 0.03) |

Estimates from population and within-sibship models between educational attainment and low-density lipoprotein cholesterol concentration from HUNT, UK Biobank, fixed-effects meta-analysis of HUNT and UK Biobank, and the Within Family Consortium.

Abbreviations: CI = confidence interval, MR = Mendelian randomisation, SD = standard deviation.

**Table S2. Educational attainment and high-density lipoprotein cholesterol concentration**

| **Model** | | **Change in outcome per SD increase in educational attainment (SD, 95% CI)** | | |
|---|---|---|---|---|
| | | HUNT | UK Biobank | Combined |
| Observational model | Population | 0.03 (0.02 to 0.04) | 0.03 (0.03 to 0.04) | 0.03 (0.02 to 0.04) |
| | Within-sibship | 0.02 (0.00 to 0.03) | 0.00 (0.00 to 0.01) | 0.01 (0.00 to 0.02) |
| 1-sample MR | Population | 0.25 (0.16 to 0.35) | 0.10 (0.08 to 0.13) | 0.18 (0.13 to 0.23) |
| | Within-sibship | 0.38 (0.16 to 0.60) | 0.11 (0.01 to 0.21) | 0.25 (0.13 to 0.37) |
| | | | | Consortium |
| 2-sample MR | Population | | | 0.26 (0.21 to 0.32) |
| | Within-sibship | | | 0.18 (0.09 to 0.27) |

Estimates from population and within-sibship models between educational attainment and high-density lipoprotein cholesterol concentration from HUNT, UK Biobank, fixed-effects meta-analysis of HUNT and UK Biobank, and the Within Family Consortium.

Abbreviations: CI = confidence interval, MR = Mendelian randomisation, SD = standard deviation.

**Table S3. Educational attainment and trigylcerides concentration**

| **Model** | | **Change in outcome per SD increase in educational attainment (SD, 95% CI)** | | |
|---|---|---|---|---|
| | | HUNT | UK Biobank | Combined |
| Observational model | Population | –0.08 (–0.09 to –0.07) | –0.07 (–0.08 to –0.05) | –0.07 (–0.08 to –0.07) |
| | Within-sibship | –0.06 (–0.08 to –0.04) | –0.02 (–0.03 to 0.00) | –0.04 (–0.05 to –0.03) |
| 1-sample MR | Population | –0.22 (–0.31 to –0.13) | –0.29 (–0.37 to –0.22) | –0.26 (–0.32 to –0.20) |
| | Within-sibship | –0.38 (–0.61 to –0.15) | –0.41 (–0.72 to –0.10) | –0.39 (–0.59 to –0.20) |
| | | | | Consortium |
| 2-sample MR | Population | | | –0.26 (–0.32 to –0.21) |
| | Within-sibship | | | –0.11 (–0.21 to –0.01) |

Estimates from population and within-sibship models between educational attainment and triglycerides concentration from HUNT, UK Biobank, fixed-effects meta-analysis of HUNT and UK Biobank, and the Within Family Consortium.

Abbreviations: CI = confidence interval, MR = Mendelian randomisation, SD = standard deviation.

**Table S4. Educational attainment and cardiovascular disease risk**

| **Model** | | **Change in outcome per SD increase in educational attainment (HR, 95% CI)** | | |
|---|---|---|---|---|
| | | HUNT | UK Biobank | Combined |
| Observational model | Population | 0.91 (0.88 to 0.94) | 0.85 (0.80 to 0.89) | 0.88 (0.85 to 0.91) |
| | Within-sibship | 0.93 (0.88 to 0.98) | 0.96 (0.87 to 1.05) | 0.94 (0.89 to 1.00) |
| 1-sample MR | Population | 0.58 (0.46 to 0.74) | 0.69 (0.50 to 0.97) | 0.63 (0.52 to 0.78) |
| | Within-sibship | 0.29 (0.15 to 0.55) | 0.65 (0.14 to 3.01) | 0.43 (0.19 to 0.97) |
| | | | | Consortium |
| 2-sample MR (OR) | Population | | | 0.94 (0.92 to 0.96) |
| | Within-sibship | | | 0.96 (0.91 to 1.01) |

Estimates from population and within-sibship models between educational attainment and cardiovascular disease from HUNT, UK Biobank, fixed-effects meta-analysis of HUNT and UK Biobank, and the Within Family Consortium.

Abbreviations: CI = confidence interval, HR = hazard ratio, MR = Mendelian randomisation, OR = odds ratio, SD = standard deviation.

**Table S5. Educational attainment polygenic score and five outcomes**

| | | Change in outcome per SD increase in educational attainment PGS (SD, 95% CI) | | |
|---|---|---|---|---|
| | | HUNT | UK Biobank | Combined |
| Education | Population | 0.13 (0.12 to 0.14) | 0.16 (0.15 to 0.18) | 0.15 (0.14 to 0.16) |
| | Within-sibship | 0.08 (0.07 to 0.10) | 0.07 (0.05 to 0.09) | 0.08 (0.06 to 0.09) |
| LDL-C | Population | –0.02 (–0.03 to –0.01) | –0.01 (–0.02 to 0.00) | –0.02 (–0.03 to –0.01) |
| | Within-sibship | 0.00 (–0.02 to 0.02) | 0.00 (–0.02 to 0.02) | 0.00 (–0.01 to 0.01) |
| HDL-C | Population | 0.03 (0.02 to 0.05) | 0.02 (0.01 to 0.02) | 0.03 (0.02 to 0.03) |
| | Within-sibship | 0.03 (0.01 to 0.05) | 0.01 (0.00 to 0.01) | 0.02 (0.01 to 0.03) |
| TG | Population | –0.03 (–0.04 to –0.02) | –0.05 (–0.06 to –0.04) | –0.04 (–0.05 to –0.03) |
| | Within-sibship | –0.03 (–0.05 to –0.01) | –0.03 (–0.05 to –0.01) | –0.03 (–0.04 to –0.02) |
| CVD (HR) | Population | 0.93 (0.90 to 0.96) | 0.94 (0.89 to 0.99) | 0.94 (0.91 to 0.97) |
| | Within-sibship | 0.90 (0.85 to 0.95) | 0.97 (0.87 to 1.08) | 0.93 (0.88 to 0.99) |

Estimates from population and within-sibship models between an educational attainment polygenic score and five outcomes from HUNT, UK Biobank, and fixed-effects meta-analysis of HUNT and UK Biobank.

Abbreviations: CI = confidence interval, CVD = cardiovascular disease, HDL-C = high-density lipoprotein cholesterol, HR = hazard ratio, LDL-C = low-density lipoprotein cholesterol, MR = Mendelian randomisation, OR = odds ratio, PGS = polygenic score, SD = standard deviation, TG = triglycerides.
